# Size and duration of COVID-19 clusters go along with a high SARS-CoV-2 viral load : a spatio-temporal investigation in Vaud state, Switzerland

**DOI:** 10.1101/2021.02.16.21251641

**Authors:** Anaïs Ladoy, Onya Opota, Pierre-Nicolas Carron, Idris Guessous, Séverine Vuilleumier, Stéphane Joost, Gilbert Greub

**Author notes:** Equally contributed (Séverine Vuilleumier, Stéphane Joost, Gilbert Greub).

## Abstract

To understand the geographical and temporal spread of SARS-CoV-2 during the first wave of infection documented in the canton of Vaud, Switzerland, we analysed clusters of positive cases using the precise place of residence of 33’651 individuals tested (RT-PCR) between January 10 and June 30, 2020. We identified both space-time (SaTScan) and transmission (MST-DBSCAN) clusters; we estimated their duration, their transmission behavior (emergence, growth, reduction, etc.) and relative risk. For each cluster, we computed the within number of individuals, their median age and viral load.

Among 1’684 space-time clusters identified, 457 (27.1%) were significant (p ≤ 0.05), i.e. harboring a higher relative risk of infection, as compared to other regions. They lasted a median of 11 days (IQR 7-13) and included a median of 12 individuals per cluster (IQR 5-20). The majority of significant clusters (n=260; 56.9 %) had at least one person with an extremely high viral load (above 1 billion copies/ml). Those clusters were considerably larger (median of 17 infected individuals, p < 0.001) than clusters with subjects showing a viral load lower than 1 million copies/ml (median of 3 infected individuals). The highest viral loads were found in clusters with the lowest average age, while clusters with the highest average age had low to middle viral load. Interestingly, in 20 significant clusters the viral load of three first cases were all below 100’000 copies/ml suggesting that subjects with less than 100’000 copies/ml may still have been contagious. Noteworthy, the dynamics of transmission clusters made it possible to identify three diffusion zones, which mainly differentiated rural from urban areas, the latter being more prone to last and spread in a new nearby clusters.

The use of geographic information is key for public health decision makers to mitigate the spread of the virus. This study suggests that early localization of clusters help implementing targeted protective measures limiting the spread of the SARS-CoV-2 virus.

## Introduction

The novel coronavirus SARS-CoV-2 causing the COVID-19 disease has impacted our societies on an unprecedented scale. The number of infected people increased rapidly globally, with more than 84 million confirmed cases as of January 2021 and more than 1.8 million deaths (1, 2). The quick spread of the disease has challenged international experts and policymakers to implement strategies according to the local virus spread, healthcare resources, economic and political factors (Nicola et al., 2020; see also the cross-country analysis of COVID-19 response: https://analysis.covid19healthsystem.org/). Around the globe, tracing, lockdowns and quarantines have been implemented to contain the spread and impacted more than four billion people worldwide (4). These measures aim to protect the significant fraction(~22%) of the world population at risk of severe COVID-19 (5). They raise major challenges related to their dramatic impact on individual health, the capacities of our healthcare system and on our economy (4, 6, 7).

COVID-19 outbreaks occur by spreading via close contact forming clusters of cases. A critical challenge to contain the spread of the virus lies (i) in the early detection of these clusters, which reflects active viral transmission (8) and (ii) in the understanding of their spatial and temporal evolution (9). Geospatial tools using the precise location of the place of residence of tested individuals are highly effective to oversee an epidemic (10, 11). They allow for implementation of strategies to control the local disease spread in space and time (12).

Although widely used, there is no general agreement on the definition and the concepts of cluster, outbreak and hotspot, and more specifically in a spatial context. The information available from public health departments around the world globally converge, despite differences exist. The term “cluster” generally refers to a temporal aggregation and a spatial concentration of infections cases. COVID-19 clusters are constituted of two or more test-confirmed cases – three or more in France (www.santepubliquefrance.fr) and Switzerland, even 10 or more in New Zealand (www.health.govt.nz) – among individuals associated with a specific non-residential setting with illness onset dates within 7 to 14 days. To further label clusters as outbreak, one must also either 1) have identified direct exposure between at least two of the test-confirmed cases in that setting (for example under one meter face to face) and this during the infectious period of one of the cases, or 2) when there is no sustained local community transmission, one must have noticed the absence of an alternative source of infection outside the setting for the initially identified positive cases (13). Clusters are also assimilated to the concept of “hotspot”, which is not clearly defined neither but often used in spatial epidemiology (14). The World Health Organization (WHO) defined a set of methods and procedures to identify epidemic hotspots and to use them for global surveillance of a population (UNAIDS/WHO, 2013). Beside, infectious diseases studies have proposed methods to identify spatial clusters and characterize them (16, 17).

The identification of areas of high prevalence of any phenomenon constitutes a specific research domain in spatial statistics. Point pattern analysis (18) and local spatial autocorrelation methods were applied for the detection of disease clusters (19). In the current COVID-19 pandemic, Zhang et al. (2020) used local Moran’s statistics to identify clusters in China, but at a large geographic scale and using incident cases aggregated at the level of large administrative units. Among studies involving geospatial information reviewed by Franch-Pardo et al. (2020), few characterized the spread of COVID-19 in space and time (e.g. Desjardins et al., 2020) and even less used spatial statistics to detect clusters at a local scale (22, 8). More studies on local and regional scales that consider demographic characteristics of a population at risk are needed to provide timely information to enable accurate prevention and containment measures (10). Indeed, the precise detection of spatial clusters, the description of their dynamics and evolution over time in a geographic context are key to inform decision-makers, to deploy smart testing overtime and to provide targeted health and prevention interventions at a local scale (23).

The persistence in time of clusters was shown to be associated with socio-economic deprivation (22), but their size and duration are also likely to be due to so-called “super-spreader” individuals or events (24). They relate to the evidence for large variation in individual reproductive number (25). A super-spreader is considered to greatly contribute to the transmission of an infectious disease. Stein (2011) estimates that it would correspond to a 20/80, i.e. 20% of individuals for up to 80% of the transmission. Super-spreaders exist among the SARS-CoV-2 infected persons (27); they are more likely to be highly infectious, a mechanism suggested to be related to high viral loads (28). Noteworthy, as shown recently (29), viral load found in SARS-CoV-2 infected people appears to be similar to that observed with other respiratory viruses such as influenza B and to be similar across ages. Why SARS-CoV-2 exhibits such a high reproductive number (R0) of about 2 to 3.5 (30), and whether transmission pattern, cluster duration and size correlate somehow with viral load remains to be explored in a detailed spatio-temporal context.

Here, we characterized the spatial and temporal dynamics of the first wave of SARS-CoV-2 infection in the canton of Vaud (western Switzerland) through the detection and the location of clusters, as well as of their characteristics such as size, duration and composition (number and age of individuals and their viral load). We used the results of the SARS-CoV-2 RT-PCR tests (n= 33’651) performed by the Microbiology Laboratory of the Lausanne University Hospital (CHUV) between January 10 and June 30, 2020 (with a first positive case on 2^nd^ March). The data collected are results of RT-PCR tests, viral load (copies/ml) when the test is positive, age and geographic location of the address of residence of individuals tested. We used on the one hand a spatial scan approach (31, 32) (i) to detect spatio-temporal clusters of COVID-19 on a daily basis, (ii) to disentangle the relationships between cluster size, duration and composition, and (iii) to assess the importance of viral load in the evolution of the clusters. On the other hand, we implemented the Modified Space-Time DBSCAN (MST-DBSCAN) algorithm (33) to characterize the diffusion dynamics of transmission clusters. Finally, we discussed the effect of a soft lockdown such as deployed in Switzerland from March 19 to April 27, 2020, on the dynamics of the spread of the virus.

## Material and Methods

### Patients

Patients exhibiting symptoms compatible with COVID-19, such as fever, cough, dyspnea, smell loss or taste loss were tested by RT-PCR for the presence of the SARS-CoV-2 in their nasopharyngeal secretions, at least when considered vulnerable (e.g. with immunosuppression, obesity, chronic obstructive lung disease or age > 65 years) or when likely exposed to vulnerable cases (e.g. healthcare workers or subject living with vulnerable persons). Contacts of most positive cases were also tested, even when asymptomatic, in order to define need of a 10 days quarantine period or isolation. Precise address of residence was prospectively collected at the time of sampling as well as person age.

### SARS-COV-2 RT-PCR

Most RT-PCR were performed using the automated molecular plateform implemented at the Institute of Microbiology. It uses the Magnapure automated RNA extraction method followed by PCR amplification on QuantStudio automated systems (34) with primers described by Corman et al. (2020), later slightly modified according to Pillonel et al. (2020) to further improve PCR sensitivity. Then, from March 24, 2020, most RT-PCR were performed using the COBAS 6800 RT-PCR test, which exhibited similar performance than the home-brew automated approach (Opota et al. 2020). A few numbers of cases were tested using the GeneXpert approach to reduce time to results (38). Viral load was calculated based on the so-called “cycle threshold” (Ct), which corresponds to the number of cycles, when the fluorescent signal is above a predefined threshold (37, 38).

### Study area

All the data used were collected in the state of Vaud located in the south-west of Switzerland, north of Lake Geneva. It has an area of 3’212km2 (see Figure 6A), a population of 811’203 individuals (end of 2019), for a density of 249 inhabitants/km^2^. Of note, there are important differences in population density between the urban area of Lausanne-Morges on the shores of Lake Geneva (~3’000 inhabitants/km2), and country-side toward north where population density is of ~200 inhabitants/km2. One exception is the area of Yverdon-les-Bains directly south of the Lake of Neuchâtel with 2’200 inhabitants/km2.

### Spatio-temporal clusters

We used the SaTScan software (version 9.6.1) to detect daily space-time clusters of individuals tested positive for SARS-COV-2 in the canton of Vaud from March 2 to June 30 2020 (no positive cases between January 10 and March 2, 2020). The algorithm developed by Kulldorff (1997) tests whether a disease is randomly distributed over space and time. It uses a “moving cylinder”, with the base and height corresponding to the spatial and temporal components, respectively. Significance evaluates the excess relative risk, i.e. more than expected observed COVID-19 cases within the moving cylinder relative to randomly distributed cases over space and time. We implemented it in a daily prospective surveillance analysis. We used a discrete Poisson model, where the number of events in the geographic area (total number of positive tests) is Poisson-distributed, according to a known underlying population at risk. The spatial size of the clusters’ radius reported on maps covers a maximum of 0.5% of the total resident population (population at risk) in the canton of Vaud (N=811’203 inhabitants; SFSO, 2019). Tested individuals and the underlying population at risk were georeferenced at the centroids of a hectometric grid (40) covering the entire study area. The minimum number of positive cases considered to constitute a cluster is 3, and we restricted the temporal scanning window to a minimum of 2 days and a maximum of 14 days. The upper limit of 14 days accounts for the incubation period (generally 2 to 7 days) and for being infectious (no more than 10 days from symptoms onset). The significance of the clusters was evaluated on the basis of 999 Monte-Carlo permutations that randomize both locations (41) and times of the cases.

### Cluster evolution and diffusion zones

We used MST-DBSCAN (modified space–time density-based spatial clustering of application with noise; Kuo et al., 2018) to characterize the diffusion dynamics of clusters. MST-DBSCAN is an algorithm for detecting, characterizing, and visualizing disease cluster evolution in the geographic space and in time. It computes geographically a kernel density that considers the effect of the incubation period of an infection disease. It is based on DBSCAN (42), a non-parametric density-based clustering algorithm that groups together objects (here SARS-COV-2 positive cases) that are closely packed together (points with many nearby neighbors), marking as outliers points that lie in low-density regions. The MST-DBSCAN identifies six different cluster behaviors: a) emerge, b) grow (or increase), c) remain steady (keep), d) move, e) split or f) reduce (decrease).

We applied the MST-DBSCAN analysis to the 3’317 COVID-19 positive cases identified (among 33’651 tested individuals) and georeferenced at their precise address of residence in the canton of Vaud. Disease clusters were computed daily from March 4, 2020, to June 30, 2020. The maximum spatial radius considered was of 1’000 meters, and we have set a time window of 1 to 7 days to reflect the incubation period of the disease. A minimum number of 3 positive cases was considered to constitute a cluster. For all clusters identified, we established a typology of similar diffusion patterns in the geographical space. We associated the clusters with the postcode areas (557 units; MicroGIS, 2019) of the canton of Vaud and used them as spatial references. Then, we focused on three main cluster behaviors, which are increase (b), reduce (f) and keep (c) to characterize the diffusion type through the postcode areas. The diffusion patterns were detected using the Louvain method, a group detection algorithm using network analysis (44). This approach synthetizes the spatio-temporal information and facilitates its visualization on a single map.

## Results

### Epidemic trajectories of positive cases

A total of 33’651 subjects have been tested over a period of 6 months, of which 3’317 (9.86%) were positive by RT-PCR. Seventy-nine percent of positive cases (2609/3317) were observed between March 9 and April 5: this 4-weeks period corresponds to only 16% of the total duration of the studied period, but in this short period as many as 2’609 tests were positive (22.2% of 11’756 tests) (Figure 1A). The peak of the 1^st^ epidemic wave occurred on March 18, i.e. two days after the start of the soft lockdown implemented in Switzerland and lasting from March 16 to April 27 (see vertical dashed lines in Figure 1A). At the peak of the outbreak, as many as 180 Vaud subjects were documented positive in our laboratory in a single day (Figure 1A, dark blue). Number of positive cases decreased then considerably from May 1st. The highest proportion of positive tests was observed 4 days after the peak of the epidemic wave, with a rate of positive tests reaching 32% (Figure 1A, light blue). The rate of positive cases was relatively high at the start of the epidemic when few individuals were tested and found positive. Then the shape of percentage of positive tests followed the trajectory of the number of cases. This is likely to be related to the fact that at the beginning of the epidemics only symptomatic subjects and patient at risk were tested, and then a much wider category of individuals was tested to the point all symptomatic individuals and asymptomatic contacts could access a test.

**Figure 1.**
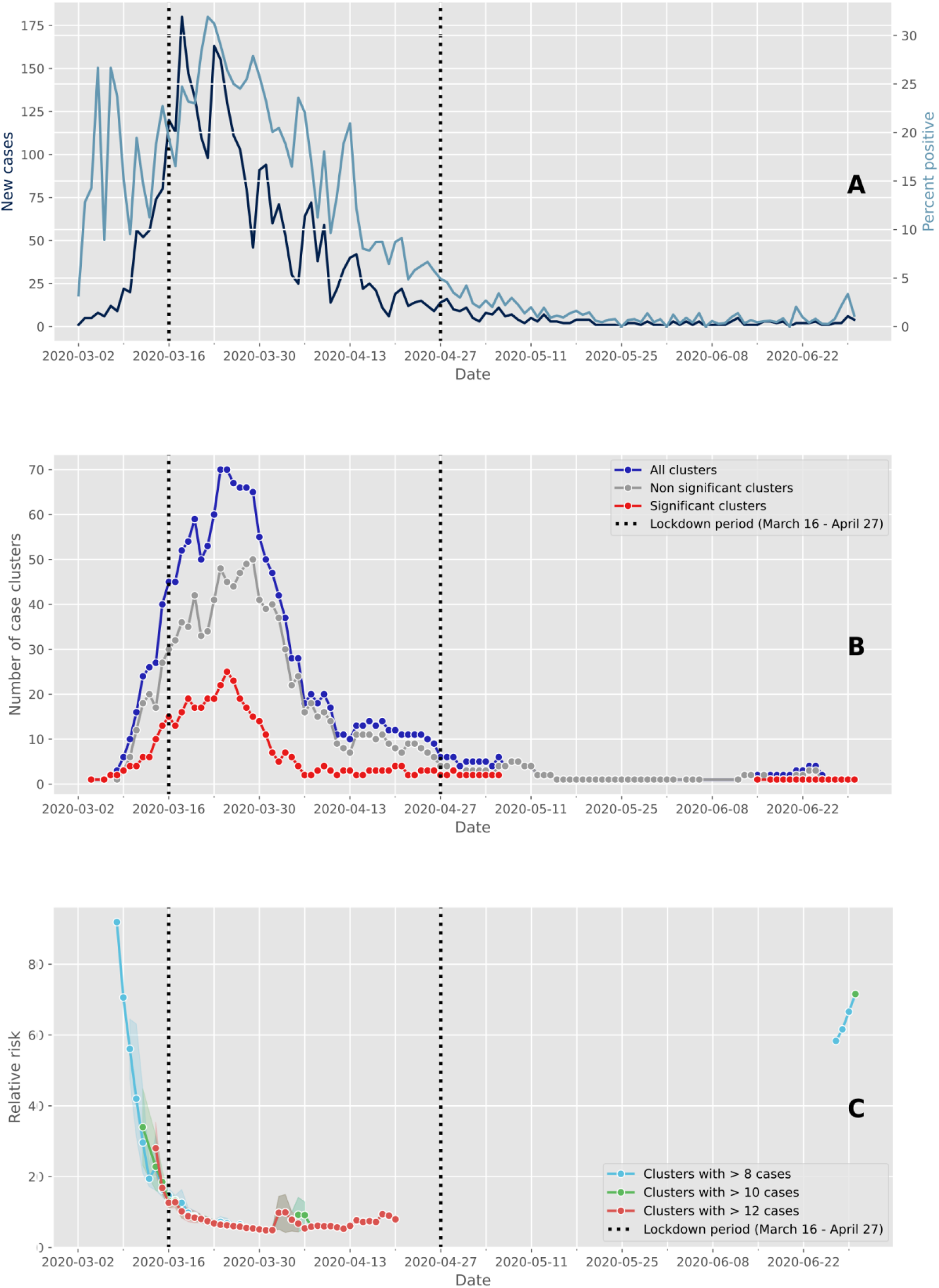
Evolution of cases and clusters through time. On the figures, the vertical dashed lines delimit the Swiss lockdown period (March 16 – April 27). (A) Epidemic trajectory of positive tests. The daily new confirmed cases are represented in dark blue whereas the percent of positive tests are shown in light blue. The peak of daily new cases occurred on March 18, while the highest proportion of positive tests was recorded on March 22. (B) Number of case clusters over time. The total number of clusters detected daily by space-time scan statistics is shown in dark blue, while the red and gray lines represent the proportion of significant clusters (p≤0.05) and non significant clusters (p>0.05) respectively. (C) Average relative risk of significant space-time clusters (p≤0.05) over time according to the within-cluster cases. As the expected number of cases was very low in some sparsely populated rural areas, resulting in extremely high relative risk values, we calculated the relative risk by considering only clusters with more than 8 (blue line), 10 (green line) and 12 cases (red line).

### Cluster detection and temporal dynamics

We identified 1’684 space-time clusters using the place of residence of patients positive to SARS-CoV-2. Among them, 457 were considered significant based on the within proportion of positive cases compared to the total documented positive cases. Highest values of both significant and non-significant clusters were observed between March 9 and April 5 (Figure 1B). Number of clusters decreased from the 1^st^ of May. Thus, the decrease in the number of positive patients following the beginning of the soft lockdown (Figure 1A) occurred about two weeks earlier than the decrease in the number of clusters (Figure 1B). Significant or not, the number of clusters displays a similar pattern through time in terms of increase and decrease but with a difference in amplitude. As shown on Figure 1C, the relative risk for new clusters was higher before the soft lockdown and about 80 days after the end of the lockdown. The size of clusters (i.e. number of cases within clusters) used to compute the relative risk does not strongly change the value of the relative risk during the core of the epidemic wave; however, it affects this value when the number of positive cases is small, i.e. at the beginning and at the end of the epidemic wave.

### Cluster composition

Significant space-time clusters generally involved a larger number of positive cases (maximum of 21 cases in average, with the largest cluster showing 43 cases on March 25) compared to non-significant ones (maximum of 11 positive cases in average; Figure 2A). Noteworthy, as shown in Figure 2A, significant clusters with more than 15 positive cases were mainly observed shortly after the soft lockdown implemented from March 16 to April 27, with one exception on April 3. Cluster durations – although limited to 14 days - increased over time from the start of the epidemic wave, showing little differences between significant and non-significant clusters. We can also observe the absence of any significant cluster from May 3 to June 16 (Figure 2B).

**Figure 2.**
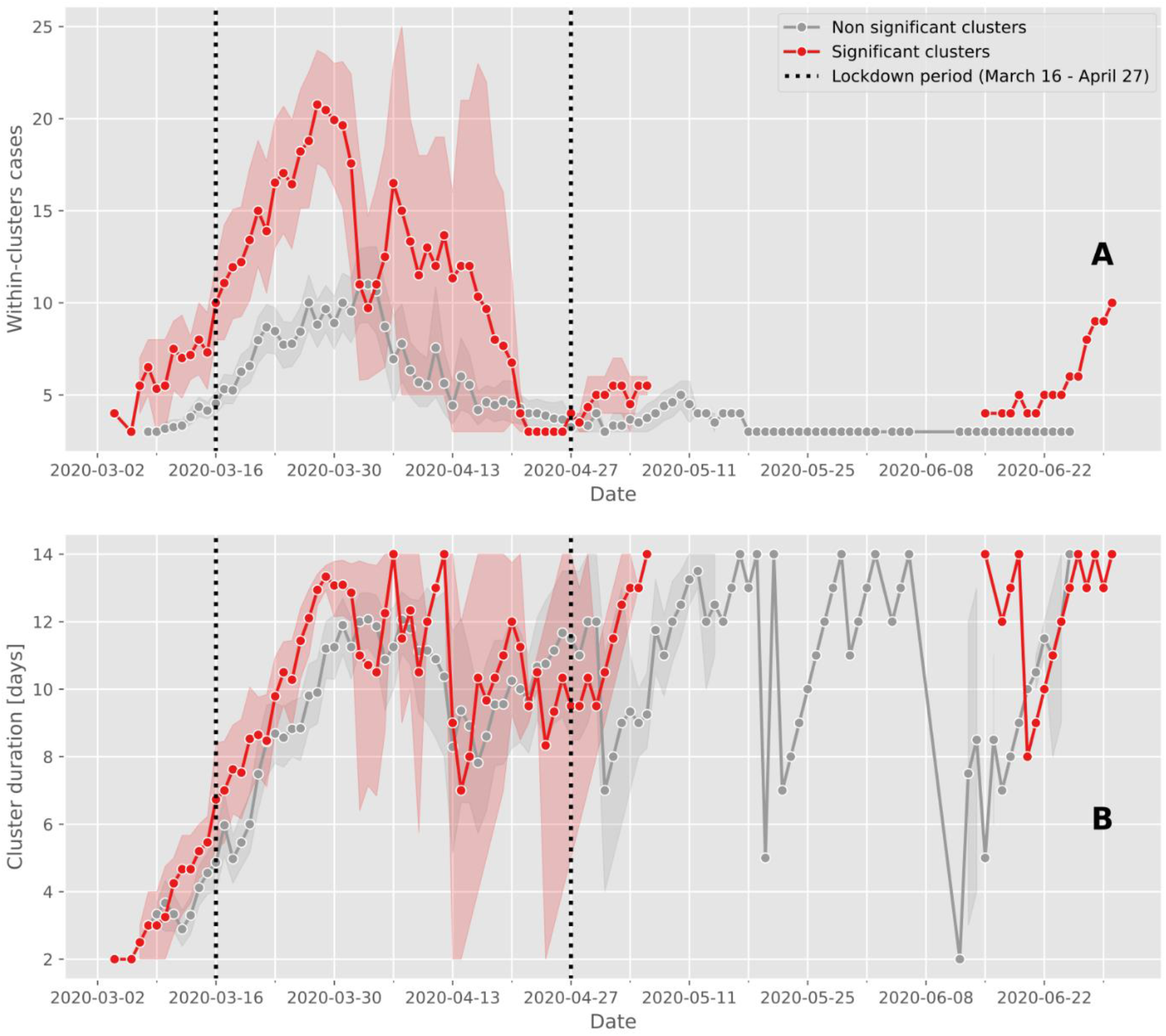
Case cluster characteristics over time. The average and confidence intervals of the number of cases within clusters (A) and the duration of clusters (B) are calculated for significant (red line) and non significant clusters (grey line). Between May 6 and June 15, the prospective space-time scan statistic detected no significant clusters. In the figures, the vertical dashed lines delimit the Swiss lockdown period (March 16 – April 27).

### Viral load in clusters

Clusters were defined by the presence of at least 3 positive cases within a limited geographic area, as documented in the SaTScan analysis. All clusters were then characterized according to the nasopharyngeal viral load of the cases documented in each cluster (Table 1). Significant clusters can have patients with viral load as low as those found in non-significant clusters, i.e. even below 10’000 copies/ml (Supp. Mat. 3). Thus, 5 significant clusters were composed of 3 cases exhibiting a viral load below 10’000 copies/ml at time of testing. However, significant clusters were more likely to be detected when viral loads were above 100 million copies/ml (Figure 3). Finally, as many as 18 significant clusters with at least 1 subject showing between 1 billion and 10 billion copies/ml were documented on March 24 (Figure 3, pink curve). The frequency distribution of viral load in significant clusters significantly differs from the distribution of viral load in non-significant clusters and outside clusters (Kolmogorov–Smirnov test, two-sample case, p<0.001, see Supp. figures 1 and 2).

**Table 1.** Classification of the space-time clusters according to the viral load of the cases involved. Within-cluster cases were identified by matching both geographically and temporally positive test subjects, geocoded at the residential address, with space-time clusters. For example, a cluster was classified as “all below 1 million” if all individuals tested positive within the cluster during its active period had a viral load below 1 million copies/ml. Noteworthy, in 20 significant clusters the viral load of three first cases were all below 100’000 copies/ml. For each cluster category, the total number of case clusters detected by prospective space-time scan statistics over the entire study period (March 2-June 20) and the proportion of significant clusters (p≤0.05) are reported.

| Case cluster viral load category | Total number of clusters | Number of significant clusters |
| --- | --- | --- |
| All below 1 million | 106 | 33 (31.13%) |
| At least one between 1 million and 10 million | 128 | 23 (17.97%) |
| At least one between 10 million and 100 million | 125 | 13 (10.40%) |
| At least one between 100 million and 1 billion | 659 | 128 (19.42%) |
| At least one between 1 billion and 10 billion | 636 | 251 (39.47%) |
| At least one above 10 billion | 30 | 9 (30.00%) |
| *Total* | 1684 | 457 |

**Figure 3.**
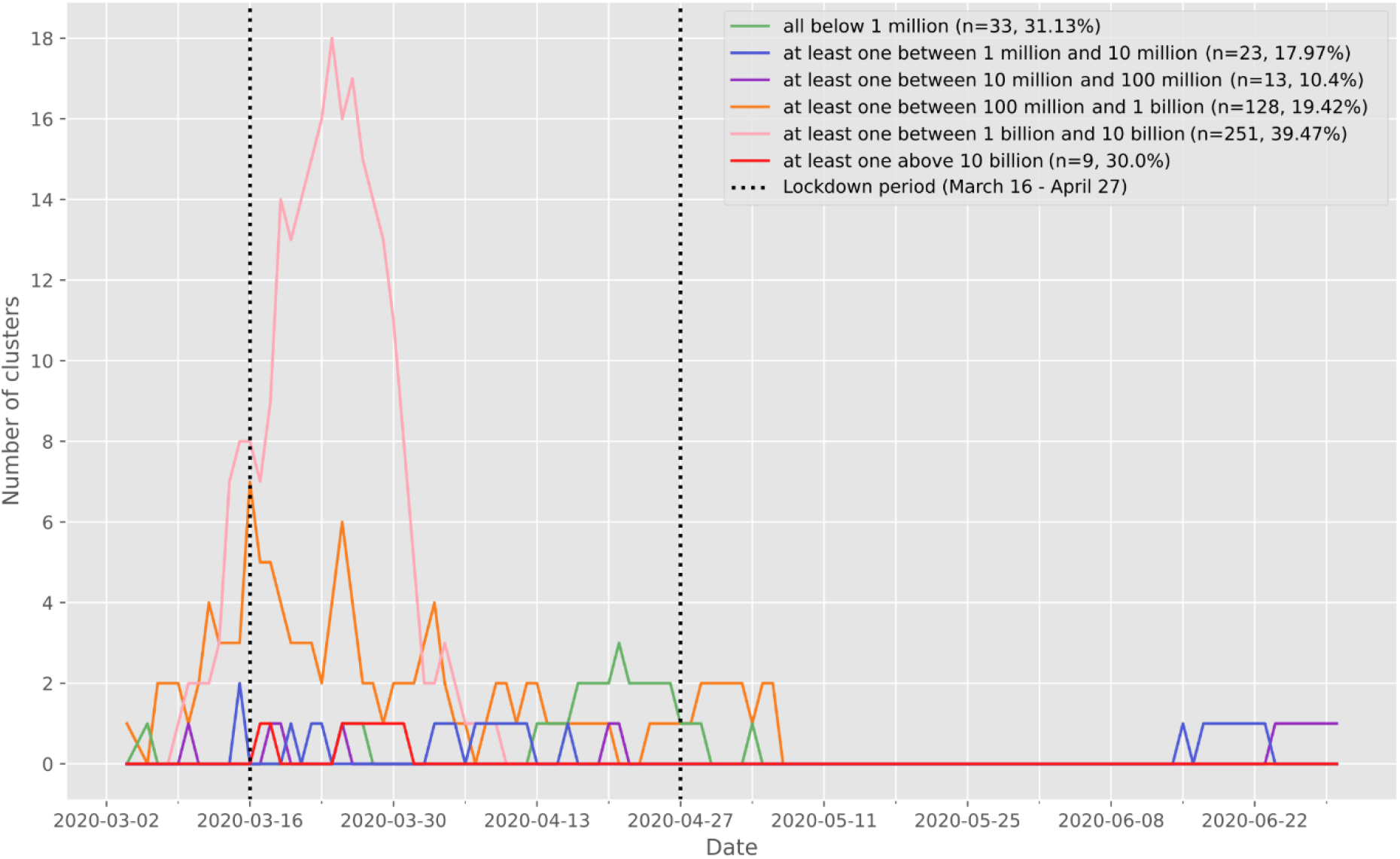
Number of significant (p≤0.05) case according to the viral load of the cases documented in each cluster. The vertical dashed lines delimit the Swiss lockdown period (March 16 – April 27). For each cluster, we extracted the positive test individuals intersecting the cluster both geographically and temporally, and we characterized the clusters according to the viral load of the individuals composing it. The clusters represented in green are therefore composed solely of individuals with a viral load of less than 1 million copies/ml. The clusters shown in blue are made up of at least one individual with a viral load between 1 million and 10 million copies/ml, and so on.

The mean viral load of the first 3 cases was also studied, in order to gain insight of the possible relationship between nasopharyngeal viral load and contagiousness, indirectly measured by the documentation of subsequent clusters. For 20 significant clusters, all first 3 cases exhibited a viral load below 100’000 copies/ml, suggesting that subjects with less than 100’000 copies/ml may still be contagious (Supp. Mat. 4). Moreover, the nasopharyngeal viral load of the first 3 cases was below 1 million copies for 40 significant clusters.

### Cluster size, duration and viral load

Cluster size (number of within cases) is positively associated with the presence of individuals with high viral load (Figure 4A). The highest viral loads measured showed a value of at least 10 billion copies/ml and occurred in the largest clusters (median number of 21 positive cases). Such a result nicely identifies super-spreading events. Even when comparing clusters harboring individuals with all viral loads below 1 million copies/ml and the ones with at least one case showing a viral load above 1 million copies/ml, the difference in cluster size was significant with a median increasing from 3 cases per cluster to 4 cases per cluster (p<0.001; figure 4A). Similar relationships were observed when considering the mean and maximal values of viral loads of the first three positive cases (Figure 5A & B).

**Figure 4.**
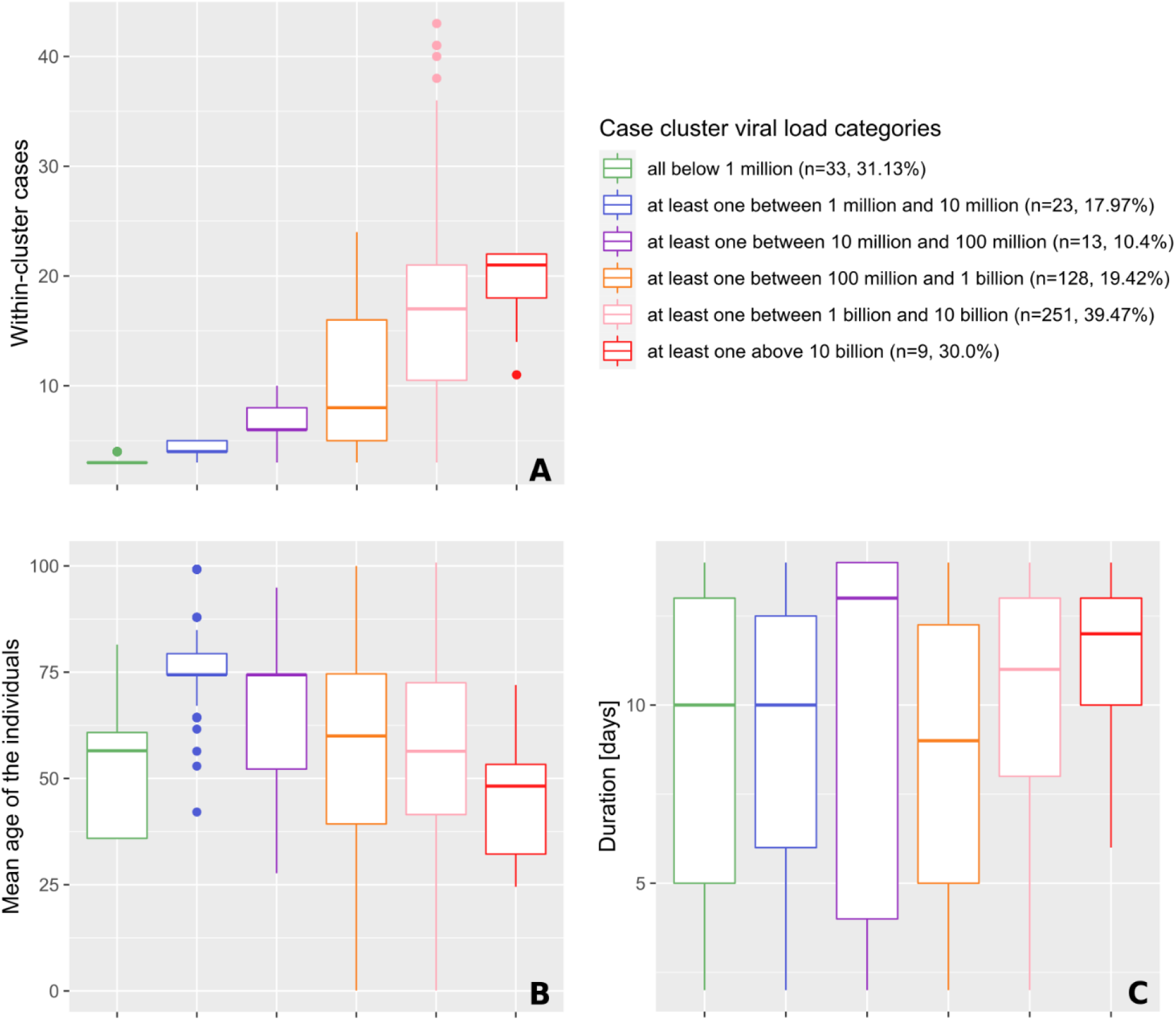
Characteristics (size, age and duration) of significant space-time clusters (p≤0.05) over the study period, categorized according to the viral load of the cases involved. The classification procedure is explained into more details in the legend of Table 1. Characteristics include the number of cases observed in clusters (A), the mean age of the positive tests individuals forming clusters (B), and the duration of clusters (C). Of note, the median number of cases was significantly higher (17 individuals) among clusters with subjects showing extremely high viral load > 1 billion copies/ml) as compared to clusters with individuals exhibiting a viral load < 1 million copies/ml (xx individuals).

**Figure 5.**
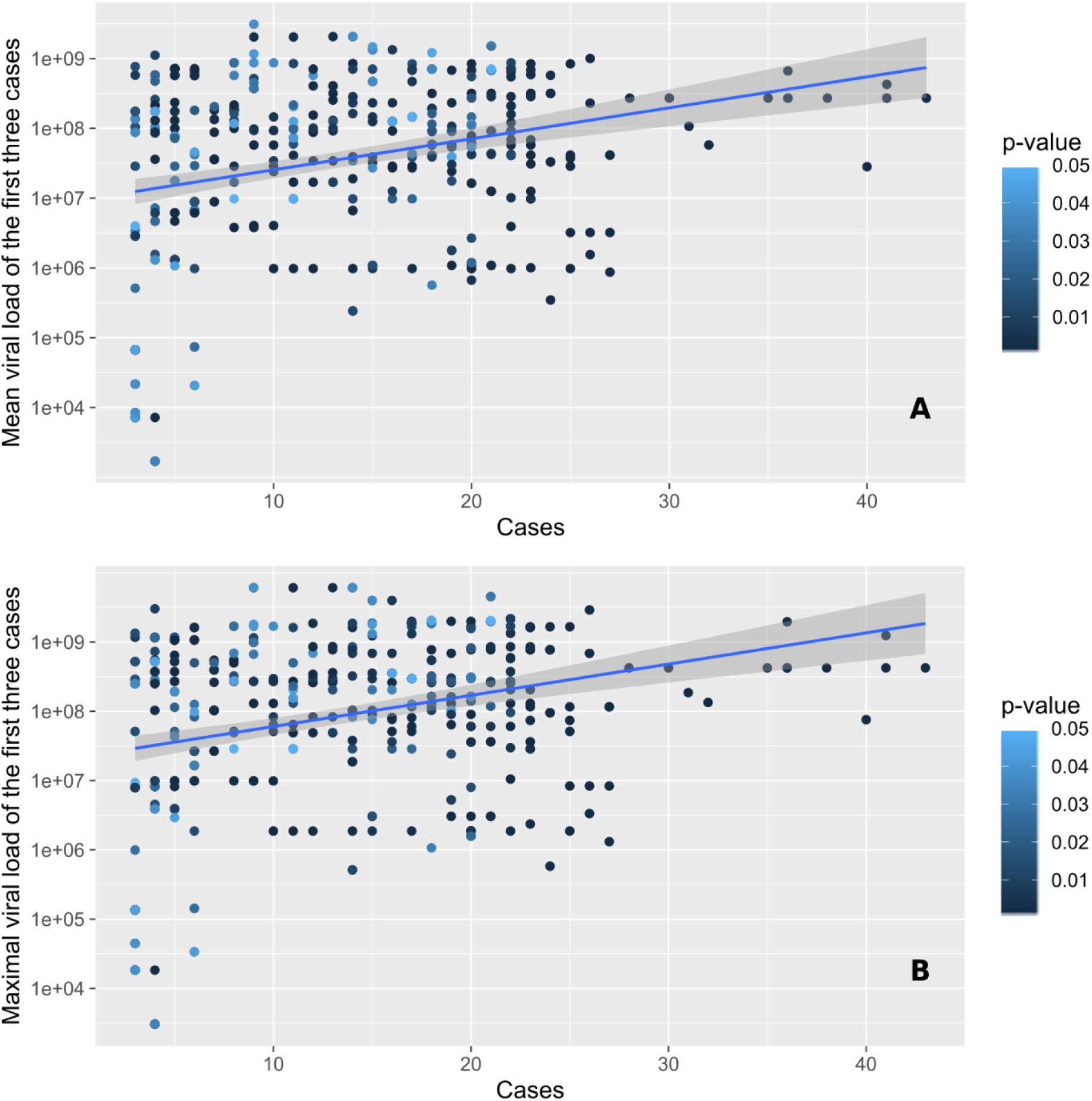
Number of cases observed within significant space-time clusters (p≤0.05) in function of the mean (A) and maximal (B) viral load of the first three cases involved. Points are colored according to the significance level of the cluster, which was assessed through 999 Monte Carlo random permutations.

Highest values of viral load were found in clusters with individuals showing the lowest average age. Clusters composed of individuals with the highest average age had low to middle viral load values (Figure 4B). Indeed, the median age of the individuals within a cluster is significantly higher when the viral load value in the cluster is between 1 and 10 million copies/ml. Then, the average age progressively decreases from 74 years to 48 years, while viral load values increase. Cluster duration is significantly different only between the orange category (100 million to 1 billion/ml) and the pink category (1 to 10 billion/ml) as clusters of the latter category last longer (mean of +0.456 days, p<0.001; see Figure 4C).

Interestingly, clusters with individuals showing the lowest average age and the highest viral load (Figure 4B) also constitute the largest clusters (Figure 4A) and those that last the longest (Figure 4C).

### Geographic distribution of the first epidemic wave

We chose six key dates to illustrate the evolution of the two types of clusters during the first wave of the epidemic in the canton of Vaud. An animation showing the spatio-temporal evolution of the clusters for the whole first wave can be visualized in Supp. Mat. 5, and in Supp. Mat. 6 for the dynamics of clusters’ behavior. Here, Figure 6 shows the spatial distribution of space-time clusters (A-F) and compares it to information translating the diffusion dynamics of the clusters (A’-F’). A detailed description of the first SARS-CoV-2 epidemic wave in the state of Vaud can be found in Box 1, illustrating the powerful and critical information that the approach offers.

**Figure 6.**
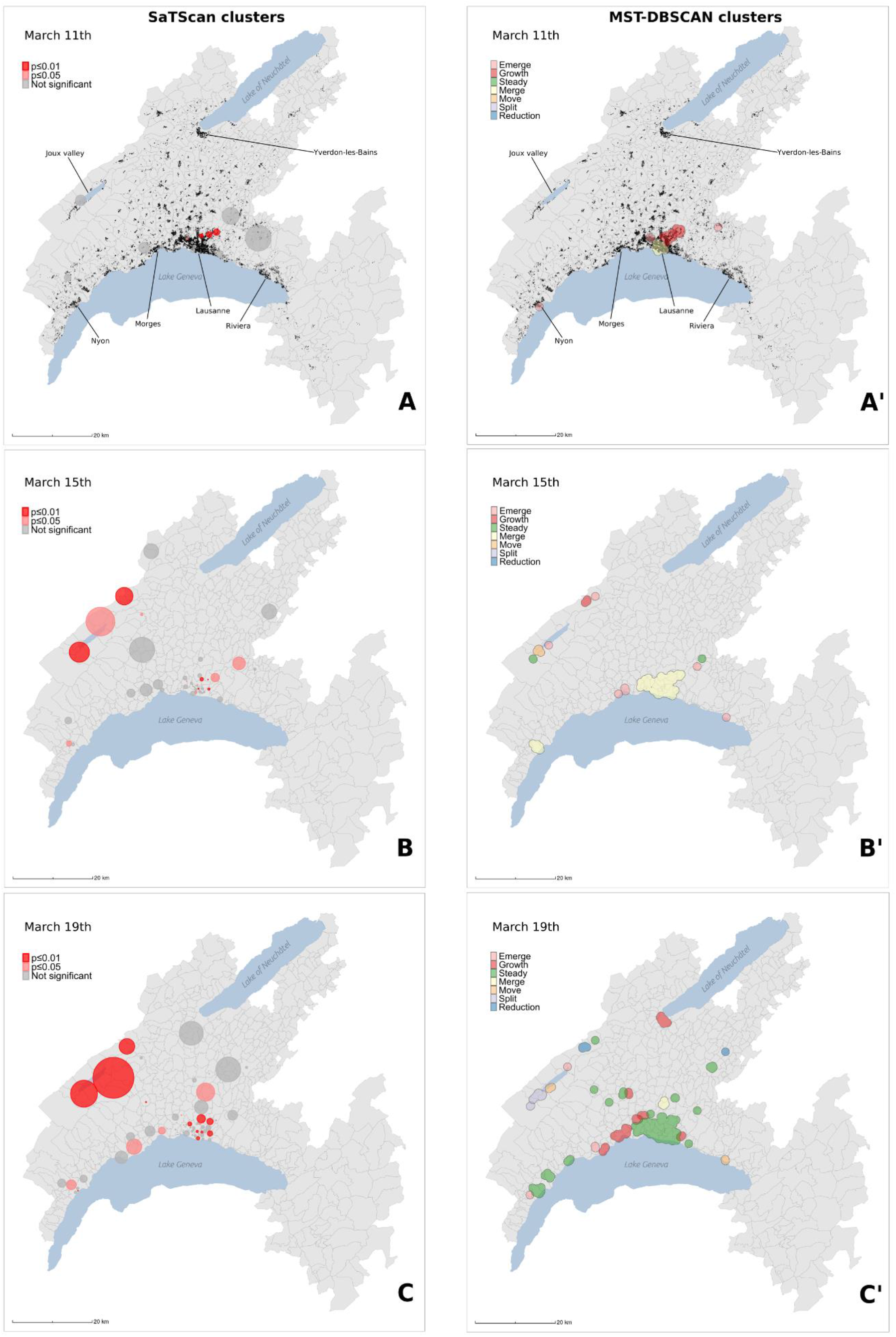
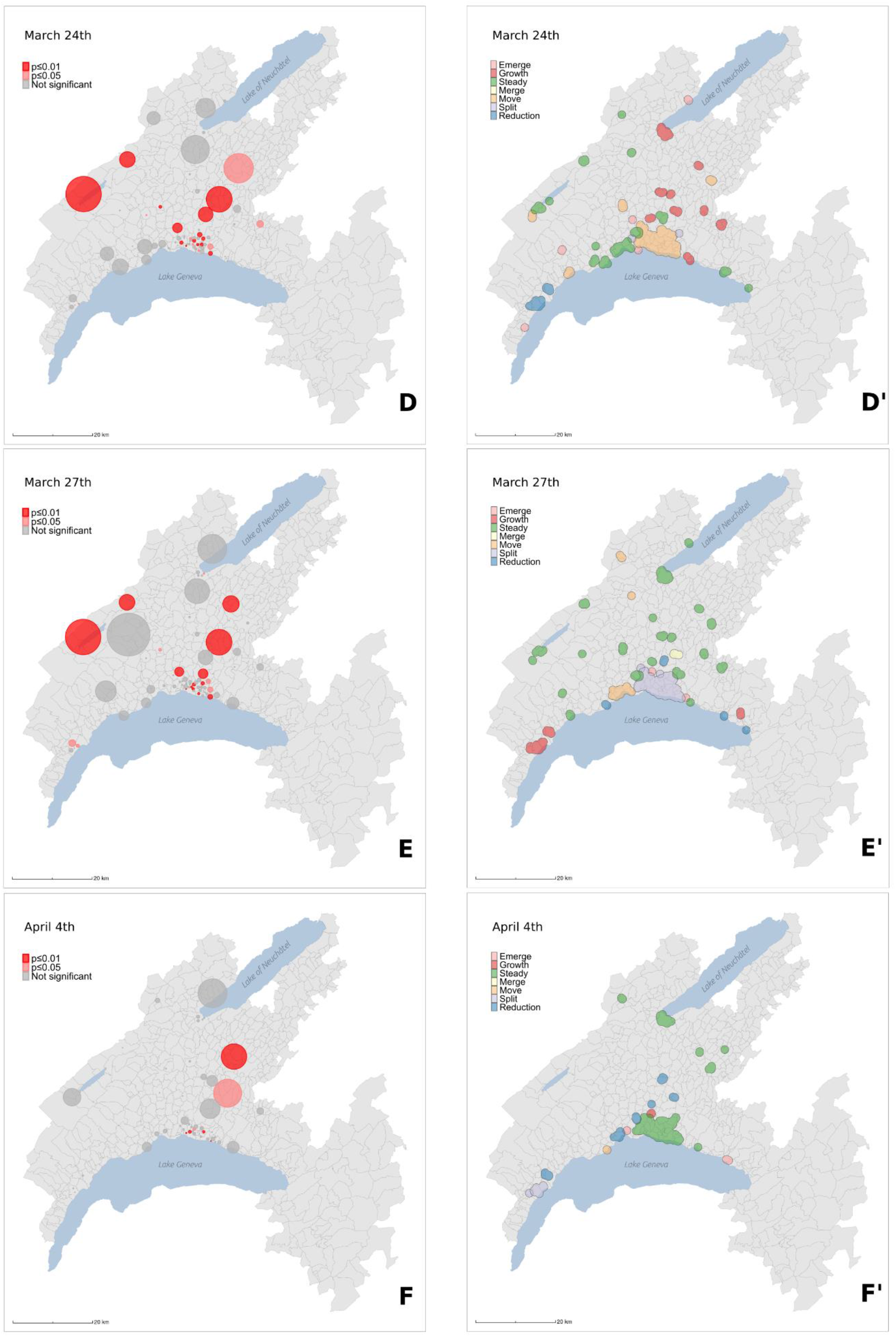
Spatial distribution of case clusters (A-F) and diffusion dynamics of transmission clusters (A’-F’) for 6 key dates (March 11, March 15, March 19, March 24, March 27, April 4) during the first epidemic wave. Case clusters resulting from the prospective Poisson space-time scan statistics (A-F) are shaded according to their significance level: dark red for statistically significant clusters with alpha = 0.01, light red for statistically significant clusters with alpha = 0.05, and grey for non significant clusters (p>0.05). Transmission clusters resulting from the MST-DBSCAN algorithm (A’-F’) are shaded according to their evolution type: emerge (pink), growth (red), steady (green), merge (yellow), move (orange), split (purple), and reduction (blue). Black points in (A) and (A’) represent the 33’651 individuals tested during the study period (January 10–June 30).

Cluster behaviors described in Box 1 were summarized with four diffusion zones shown in Figure 7A and identified at the level of postal code areas using MST-DBSCAN (Figure 7BCD). The grey diffusion zone corresponds to areas where no clusters emerged, while the green, orange and blue diffusion zones differ in the way clusters evolved over time. The green diffusion zones correspond to areas where the clusters immediately increased in size at the beginning of the epidemic wave (red line, Figure 7B) but decreased drastically once the soft lockdown (vertical dash line) took place. Then we observed a second peak associated with an important increase of clusters that reduced in size (red line, Figure 7B). Both red and purple curves are bimodal and tend to decrease afterwards, with a few numbers of new small peaks that plateau forming a distribution with a long right tail. Conversely, orange and blue diffusion zones show a first peak of increasing clusters later, i.e. about at the time of the start of the soft lockdown (orange & blue areas, Figure 7C and D). Both also show clusters that remain stable in size during the soft lockdown (blue line). Blue diffusion zone is the only one to show no more clusters after April 27, i.e. the end of the lockdown and that does not display a bimodal distribution of clusters. Please note that no difference in viral load was documented among these different diffusion zones (Figure 7E).

**Figure 7.**
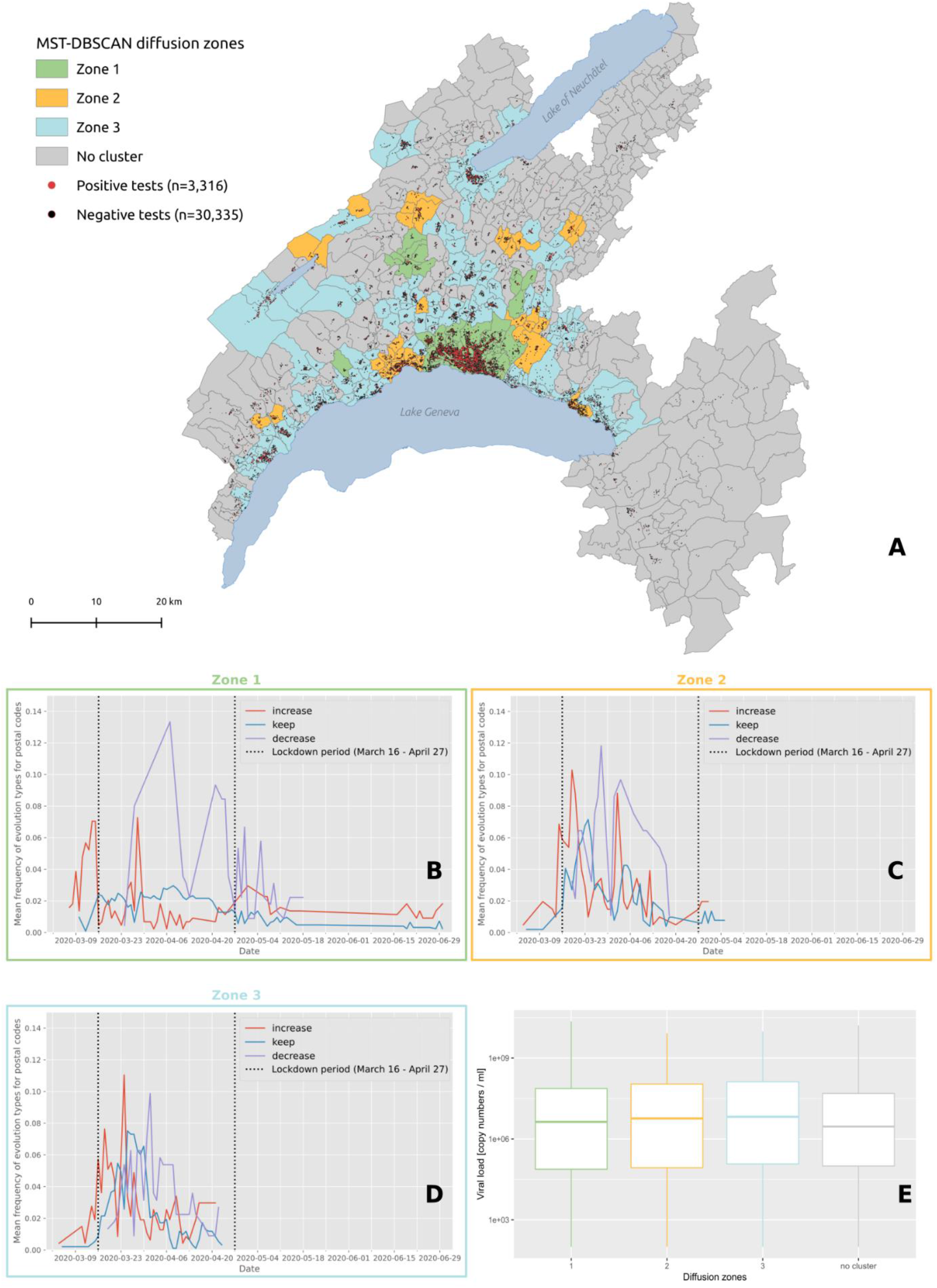
(A) Diffusion zones identified by the MST-DBSCAN algorithm. Postcode areas with the same color share similar diffusion patterns, and areas without any transmission clusters are represented in grey. The red and black dots on the map indicate the individuals tested positive (n=3’316) and negative (n=30’335) for COVID-19 in the canton of Vaud between January 10 and June 30. Below the map are shown the frequencies of major evolution types over time for zone 1 (B), zone 2 (C) and zone 3 (D). The red line corresponds to the “increase” diffusion type assigned to the transmission clusters whose area becomes larger, the blue line corresponds to the “keep” diffusion type assigned to the transmission clusters whose area remains, and the purple line corresponds to the “decrease” diffusion type assigned to the transmission clusters whose area becomes smaller. The vertical dashed lines delimit the Swiss lockdown period (March 16 – April 27). (E) Distribution of the viral load of test-confirmed cases living in each diffusion zone.

## Discussion

The discussion is divided into three majors part. The first highlights results that uncover new information on COVID-19 clusters, the second explicits limitations in the interpretation of the results, the third presents added value of the methods used to tackle epidemics and to evaluate the effect of lockdown strategies.

## New information on COVID-19 clusters

### A temporal lag between positive cases documentation and clusters burden

Significant clusters were mainly observed from March 15 to April 5 (red curve in Figure 1B), whereas non-significant clusters that occurred specifically in high population density areas such as Lausanne were already documented 4 to 5 days earlier and continuously occurred until mid-May (grey curve on Figure 1B, and Figure 6A). We observed a time shift between the decrease of the number of positive cases and the decrease of the number of clusters. This delay could be explained by the fact that most positive cases might have been at the origin of lasting clusters, i.e. lasting more than 10 days starting at the time when positive cases are identified. Interestingly, the number of patients hospitalized at Lausanne University Hospital (CHUV) and the number of deaths due to COVID-19 in the Canton of Vaud also followed the same epidemic curve, but with a 2 weeks delay (personal communication, G. Greub).

### Viral load is strongly informative on the presence and size of SARS-CoV-2 clusters

Our results show that clusters at the peak of the SARS-CoV-2 epidemic wave are composed of individuals showing a high viral load. Cluster size is positively associated with the presence of individuals with a high viral load among the significant clusters, although 40 clusters had their 3 first cases exhibiting a viral load below 1 million copies/ml, including 33 clusters that all had their cases with a nasopharyngeal viral load below 1 million copies. Moreover, as many as 20 clusters were composed of cases that initially all had a viral load below 100’000 copies/ml, suggesting that subjects with less than 100’000 copies/ml may still have been contagious.

The fact that significant clusters are composed of patients with viral load as low as those found in non-significant clusters further supports the hypothesis of community transmission with low level of viral load. Nevertheless, this may also reflect a statistic bias since large clusters with more than 10 individuals are more likely to get at least one individual with a very high viral load.

### Advantage of RT-PCRs over antigen-based testing

Given the relatively low sensitivity of antigen tests, if we had used these assays, about 24 clusters would have been missed or identified late, when the first 3 cases would have led to additional cases. Indeed, the 20 significant clusters with a viral load of the three first cases below 100’000 copies/ml would not have been detected with antigen tests, given their limit of detection (for the best ones) of about 100 to 200’0000 copies/Ml (45). Moreover, the clusters with a case between 100’000 copies/ml and 1 million copies/ml will also not have been detected in 5% of cases given an overall antigen sensitivity for such viral load of about 80% (45). Thus, altogether, an antigen-based strategy would miss about 5% (24/457) of the significant clusters.

### High viral load in large clusters with the youngest group age

Within clusters we found a clear negative relationship between age and level of viral load measured (Figure 4B), and between cluster size and viral load (Figure 4A). Indeed, while a high viral load was found in large clusters with the youngest group age, low to intermediate viral load was measured in small clusters constituted of older group age. This suggests that large clusters were generated by active individuals belonging to the working population and that super-spreader events might be at the origin of such large clusters. Surprisingly, when the level of viral load is analyzed across age classes, no relationship was documented (Supp. Mat. 7), meaning that useful information emerges within clusters. Indeed, the characterization of the clusters provides a deeper analysis of the mechanisms behind the progression of an epidemic and the geographic analysis of clusters of cases might constitute a type of investigations to favor in the future.

### Non-significant clusters also convey information on the progression of the epidemic

Significant and non-significant clusters both show the same epidemiological trajectories. Indeed, they display similar patterns in terms of increase and decrease in size with a difference in amplitude only. This suggests that the occurrence of clusters, even non-significant, is a good estimator of the epidemic situation. However, significant and non-significant clusters differ in terms of number of cases and measured viral load, but not duration. This suggests (i) that non-significant clusters might correspond to transmission events unrelated to subjects with very high viral load, (ii) that they translate a lower impact on the population in terms of viral spread, and (iii) that they express a transition towards or from a significant spatio-temporal configuration.

## Limitations

### Tested population is not homogeneous through time

During the course of the studied epidemic wave, recommendations for testing requested by the authorities have regularly changed. Initially only symptomatic patients at risk and health workers were tested. Then, since mid-March, a wider portion of the population was progressively tested, although younger subjects still were reluctant to get tested. This might have generated heterogeneity in our longitudinal investigation. Moreover, tests could have been done at different stages of the infection (early, late, etc.) and not be representative of the right window of infection. Days of the week might also generate differences in number of positive tests as the number of tests was often reduced over the weekend, for example some persons preferred to be tested only on the next Monday, to avoid quarantine over the week-end.

### Positive cases might be missing

Our estimate of the number of positive cases might not be fully representative of the epidemics, particularly at the beginning of the event when only symptomatic patients at risk and health workers were tested. We might assume that their close relatives might also have been infected but were not tested at that time due to reagents shortage. This bias might however have a limited impact on the assessment of cluster size, since any person in contact with the positive cases (documented by the contact tracing team) was tested. Besides, a source of underestimation may be the false negative RT-PCRs, due to imperfect nasopharyngeal sampling. They might however not be of importance as the clinical sensitivity of RT-PCR performed on nasopharyngeal samples in our laboratory is very good, i.e. about 96 to 98% (46, 47). Similarly, the rate of false positives in the same laboratory is estimated to be lower than 1/10’000 tests, thank to full automation and bar-coding that have been settled to prevent human errors and samples/tubes inversion (34). Finally, missing cases might be due to the fact that that some people living or working at the border of the state might have been tested in another state, but again this seems to have a limited impact since more than 80% of all samples tested for SARS-CoV-2 in 2020 have been taken from subjects living in the canton of Vaud.

## Added value of the methods used

### Geographic clusters to characterize the epidemics: a key tool for intervention

Beyond the fact that a formal definition of what a cluster is in a geographical context is lacking, the statistical approaches used make implicit assumptions that – through different parameters – have a direct influence on cluster detection and on how to interpret them. We used two complementary approaches that highlight different key aspects of disease clustering. Space-time scan statistics detect the geographical location of case clusters, assess their significance, and characterize their relative risk and duration. This prospective approach is particularly appropriate for the establishment of a daily surveillance system, since it identifies ‘alive’ clusters only, i.e. having an excess of relative risk on the day of analysis (48). Unlike other detection methods, this approach search for clusters without imposing the specification of their size and allow for analysis of area with heterogeneous population densities. Indeed it identifies a cluster if risk of disease within a space-time cylinder (radius = space, and height = time) is higher than outside. This type of information is key for public health authorities to target neighborhoods and calibrate protective or preventive measures to be deployed.

As for the MST-DBSCAN algorithm, it characterizes the diffusion dynamic of the transmission clusters. Here the input parameters require a precise definition of the incubation period, the cluster transmission areas, and a minimum number of spatio-temporal neighbors required to form a cluster (33). The output is a cluster typology according to their behaviors, which is of interest to design sets of appropriate measures to control them. The space-time (SaTScan) and the diffusion-type (MST-DBSCAN) analyses thus provide complementary results in terms of clusters emergence, duration, and demographic characteristics. The two approaches used in conjunction, allow thus for detailed monitoring of the disease’s epidemic trajectory and populations at risk and offer adequate tools for governments to both prioritize interventions on excess-risk locations and develop adapted strategies to control cluster diffusion types.

### Maps reflect the chronology of the epidemic

The results displayed on static and animated maps well reflect the chronology of the sanitary situation during the first wave of the epidemics. For instance, the major clusters in the Joux valley area can be clearly observed on different maps (Figures 6B, 6B’, 6C, 6C’). Noteworthy, these large clusters originate from a super-spreader event that took place end of February in a religious ceremony in Mulhouse, France. Many swiss residents participated in this ceremony and additional related clusters were also observed during the same period north of the Lausanne urban area, and along the Jura mountains (e.g. Morges and Nyon). Conversely, Lausanne was early hit by clusters likely due to a first transmission event that occurred in Northern Italy.

Interestingly, the initial phase observed in the state of Vaud differ from what happened in Geneva, where the first clusters emerged in deprived neighborhoods eight days (March 5) after the first positive case (February 26) was detected (22, 8). In Vaud instead, the initial cluster was directly detected the day of the first cases (March 4), with 9 positives and in a wealthy neighborhood.

### Positive impact of soft lockdown

The soft lockdown was directly associated with a rapid reduction in the number of positive cases and this despite the increased rate of testing. The reduction takes place massively and in two clear phases in the main urban areas (see Figure 7B), while it happens as a succession of clusters increase and decrease in smaller urban centers and less dense areas (Figure 7D). However, due to the time lag between the identification of positive individuals and the constitution of clusters, the cluster burden occurred directly after the implementation of the soft lockdown. Similarly, the largest clusters, the longest duration and the clusters with individuals showing large viral loads were observed just after the same time-lag. This time lag seems shorter in urban areas as compared to rural areas, likely reflecting the faster spread of the virus in large town such as Lausanne, Morges, Nyon, Yverdon and on the Vaud Riviera. This faster spread is likely due to differences in social and cultural organization between rural and urban areas, to less available room per person in housings, with higher risk of subsequent infection in family with lower socio-economic situations.

Our results highlight the efficacy of the lockdown strategy, even soft, to control the epidemic and to decrease the number of positive cases. It also demonstrates the importance of acting when the number of positive cases increases and not waiting for the settlement of clusters. Besides, our results show that the relative risk stayed very low within all the lockdown period. Of note, the compliance of Swiss residents during the first soft lockdown is signaled by the absence of any significant cluster from May 3 to June 16. And finally, it has not escaped our notice that it is already possible to observe the beginnings of the second wave from June 22, 2020 (Figure 2A), that is to say exactly two weeks after a series of relaxations of the protective measures such as in particular the authorization of public demonstrations up to 300 people, and the opening of nightclubs (June 6, 2020).

## Conclusion

Our results highlight that cluster size goes along with the presence of individuals with high viral load, the latter being more commonly found in clusters harboring the youngest group age. This work also stresses the fact that cluster size and cluster duration are largely dependent on the viral load of a few number of individuals within a given cluster, underlying the impact of viral load on contagiousness.

Altogether, we provide robust data suggesting that transmission may occur even when all possible source cases in a cluster present a viral load lower than 100’000 copies/ml. Such low viral load cases remain undetected by antigen testing, hence underlying the importance of RT-PCRs assay in case finding and tracing strategies. This in-depth analysis suggests that even older at-risk individuals that try to avoid infection may get infected by SARS-CoV-2 and hence by one of the other cluster members, even when all cluster members exhibit a low viral load, i.e. below 100’000 copies/ml.

Finally, such a spatio-temporal characterization of clusters demonstrates the huge effect of the soft lockdown that took place in Switzerland from March 16 to April 27, 2020. Those important results have been documented thanks to the contribution of the geospatial analysis of clusters.

## Data Availability

The dataset analyzed during the current study is available from the corresponding author upon reasonable request. The dataset could not be made publicly available due to the
sensitivity of individual georeferenced SARS-CoV-2 testing data. Requests to access the data should be directed to Prof. Gilbert Greub.

https://zenodo.org/record/4541131

## Acknowledgments

Anaïs Ladoy is funded by the Direction Générale de la Santé of the canton of Vaud (DGS-Vaud) in the context of the GEOSAN project (Grant Agreement C/20-21 /037).

## Ethical statement

This study was approved by the Commission cantonale d’éthique de la recherche sur l’être humain (CER-VD), Switzerland. Authorization no. 2020-01302 (20.7.2020).

## Data availability statement

The dataset analyzed during the current study is available from the corresponding author upon reasonable request. The dataset could not be made publicly available due to the sensitivity of individual georeferenced SARS-CoV-2 testing data. Requests to access the data should be directed to Prof. Gilbert Greub.

## Author contributions

AL performed the data analyses and drafted the manuscript. AL, SV, SJ and GG conceived the study and completed the manuscript. OO, PNC, and IG participated in the design of the study and helped to draft the manuscript. All authors read and approved the final manuscript.

## Conflict of interest

The authors declare to have no conflict of interest.

## Box 1

**The first SARS-CoV-2 epidemic wave in the state of Vaud, Switzerland**

On March 11, 7 days after the first detection of a positive case (Figure 6A), we observe a phase of rapid growth and merging (see Figure 6A’) and a series of significant clusters explodes directly north of Lausanne, the main city of the state. Interestingly three out of the four clusters shown are located in wealthy areas. March 15 (Figures 6B and 6B’) is the day before the soft-lockdown. There is a multitude of clusters in the Lausanne area, among which a large fraction is significant (Figure 6B). Figure 6B’ shows that these clusters rapidly merged into a single “super cluster” deployed over the urban agglomeration. In the country-side, active clusters emerge and grow north of the lake in the Joux valley located in the Jura mountains where population density is low. Four days later, March 19 (Figures 6C and 6C’), the peak of the first wave is approaching (see Figure 1B). The number of case clusters is high in the Lausanne area (Figure 6C) but clearly stabilizes. Similar behavior is observed towards east, along Geneva lake; in the Riviera area only one moving cluster is observed (Figure 6C’), while new clusters grow in the Morges area. In the Joux valley the activity remains important, and a cluster grows in Yverdon-les-Bains, south of the Lake of Neuchâtel. On March 24 (Figures 6D and 6D’), the peak of the first wave is reached (see Figure 1B). New cases reactivate moving clusters in the center of Lausanne, while towards west the situation stabilizes and even reduces toward Geneva with no more significant cluster in the Nyon area. At the peak, a large significant cluster remains steady in the Jura, and several clusters grow in the remote, rural periphery north of the main urban area (Figures 6D and 6D’). In the north, close to the Lake of Neuchâtel, the clusters are not significant despite growing. March 27 (Figures 6E and 6E’) is the start of the important and rapid reduction phase of all clusters (see Figure 1B). The merged clusters of the Lausanne area split and most of those located on the country-side are steady. However, a significant cluster emerges in the west, in the Nyon area. On April 4 (Figures 6F and 6F’), it is the end of the peak. The Joux valley cluster is over after 25 days and clusters in the canton are either not significant anymore, or are steady, split of reduce. There is one exception north of Lausanne with a single growing cluster located in a leisure area and likely related to the presence of a school (with boarding).

## Notes

### Competing Interest Statement

The authors have declared no competing interest.

